# “You need to be able to stand up for what is right”: *MTV Shuga Naija*’s transformative impact on youth attitudes towards sexual violence in Nigeria

**DOI:** 10.1101/2023.08.23.23293994

**Authors:** Paul Hutchinson, Christopher E. Beaudoin, Dominique Meekers, Elizabeth Omoluabi, Akanni Akinyemi

**Affiliations:** Tulane University, New Orleans, Louisiana, USA; Boston University, Boston, Massachusetts, USA; Centre for Research, Evaluation Resources and Development (CRERD), Abuja, Nigeria; Obafemi Awolowo University, Ile-Ife, Nigeria Centre for Research, Evaluation Resources and Development (CRERD), Abuja, Nigeria

**Keywords:** sexual violence, stigma, youth, Nigeria

## Abstract

**Background:** In Nigeria, approximately one in ten women of reproductive age report experiences of sexual violence in the past year, with potentially enduring consequences. The impact of sexual violence can be particularly devastating for younger women and adolescents, who may face long-lasting physical, psychological, and social effects. To address this, *MTV Shuga Naija* utilizes entertainment education programming, anchored by a TV serial drama, to promote gender equality and challenge sexual violence norms and behaviors. This research examines the impact of *MTV Shuga Naija* on disclosure of sexual violence, stigma and victim-blaming attitudes, and greater dialogue about sexual violence.

**Methods:** This evaluation uses a panel survey of Nigerian youth aged 15-24 (574 females; 317 males) with data collected before and after the airing of *MTV Shuga Naija* programming. The baseline wave was conducted in person, while the endline wave was conducted via telephone due to the COVID-19 pandemic. Using the Theory of Planned Behavior, survey questions focused on norms, attitudes, beliefs, and behaviors related to sexual harassment and sexual violence, we analyze changes in self-reported sexual harassment, attitudes towards victims, and discussions with adults and family members about sexual violence for those in in *MTV Shuga Naija* program areas relative to those in two comparator states. The analysis uses a fixed-effects, doubly robust difference-in-differences (DID) estimation strategy to improve the comparability of treatment and control groups by adjusting for observed differences, thereby allowing for more precise estimation of the causal effects of MTV *Shuga Naija*.

**Results:** At baseline, the majority of both males and females across all study areas reported attitudes blaming victims rather than perpetrators for experiences of sexual violence. However, results from the doubly-robust DID models reveal significant changes in these attitudes among youth in areas exposed to *MTV Shuga Nija* programming, as well as greater disclosure of incidents of sexual violence to interviewers, possibly due to greater awareness of what constitutes sexual violence. For example, agreement with the statement, “women who wear clothes that expose their bodies are asking to be raped,” decreased by 36.6 percentage points [95% CI: -46.8pp, -26.3pp] for women and by 31.8 pp [95% CI: -45.6pp, -18.1pp] for men in areas with *MTV Shuga Naija* sexual violence programming relative to those in comparison areas. Similarly, acceptance of the attitude that "it is not rape if a woman does not fight back” declined by 28.3 pp [95% CI: -40.2pp, -16.4pp] for women and by 19.3 pp [95% CI: -35.6pp, -3.0pp] for men in treatment versus comparison areas. Contrary to hypotheses, respondents in comparison areas showed larger increases in the likelihood of talking with parents and family about sexual violence than respondents in areas targeted by *MTV Shuga Naija*, although no information was collected on the initiators, circumstances, nor content of dialogue with parents and family members, rendering difficult the interpretation of this unexpected finding.

**Conclusion:** This study provides evidence that the entertainment education approach of *MTV Shuga Naija* can indeed bring about significant progress in shifting attitudes and behaviors related to sexual violence. Even with this progress, however, victim-blaming norms and attitudes, as well as experiences of sexual violence among women -- and men -- are still widely prevalent. While *MTV Shuga Naija* has made headway in shifting the landscape surrounding this often-taboo subject in Nigeria, more work is needed to understand how communication and dialogue within families, schools, and communities, alongside greater efforts to support victims of sexual violence, can be improved and enhanced to ensure continued progress.

## Introduction

This paper analyzes data from a panel survey of Nigerian adolescents and young adults in order to estimate the impact of *MTV Shuga Naija*, an entertainment education program that aired in Nigeria from 2018 to 2020, on attitudes towards sexual violence and acknowledgement of experiences of sexual violence. The program was designed on the premise that entertainment-education can leverage the power of storytelling and media to raise awareness, change attitudes, and promote positive behavior, particularly for taboo topics and deeply ingrained norms, such as acceptance of gender-based violence (GBV).

Pervasive gender-based sexual violence exposes young women to a myriad of mental and physical challenges, warranting the development of effective solutions that address stigma, victim-blaming, and harmful gender norms. The possible post-traumatic health consequences of sexual violence are numerous both in the short and long time, including unwanted pregnancy, sexually transmitted infections, gynecological and reproductive health problems, physical trauma, cardiovascular disease, diabetes, depression, anxiety, eating disorders, emotional distress, suicidality, and subsequent engagement in potentially harmful behaviors (e.g., risky sex, smoking, alcohol, drug use) (Ellsberg et al., 2008; Fleming K., 2016; Jina & Thomas, 2013; Meekers et al., 2013; Nguyen et al., 2021). Worldwide, approximately one in three women have experienced either physical and/or sexual violence by an intimate partner or sexual violence by a non-partner at some point in their lives. Among ever-married or partnered adolescent girls, approximately one in four have experienced physical and/or sexual violence from an intimate partner, with 16 percent experiencing it in the prior 12 months (World Health Organization, 2021).

Sexual violence against women, though under-reported, is also prevalent in Nigeria. According to the 2018 Nigeria Demographic and Health Survey (NDHS), 9.1% of women aged 15 to 49 years reported that they had ever experienced sexual violence, and 4.1% reported experiences of sexual violence in the past year. Kaduna State, one of the study sites this paper, reported even higher rates; 13.8% of women aged 15 to 49 years had ever experienced sexual violence, including 7.9% in the last year (National Population Commission (NPC) [Nigeria] & ICF, 2019). The WHO estimates that about 13% of women of reproductive age in Nigeria had experienced sexual violence within the last 12 months (Sardinha et al., 2022; World Health Organization, 2021). Moreover, the 2014 Violence Against Children Survey in Nigeria indicated that the lifetime prevalence of sexual violence among youth 13 to 24 years is approximately 25% (National Population Comission (NPC) Nigeria, 2016; Nguyen et al., 2021).

Public discourse about gender-based sexual violence and encouraging the disclosure of incidents are crucial steps towards addressing the prevalence and acceptance of sexual violence, which is frequently driven by gender imbalances, male control, and inequitable gender and androcentric norms (Antai & Antai, 2008; Gage & Thomas, 2017; Pulerwitz & Barker, 2007; Umana et al., 2014; World Health Organization, 2021). However, victims’ disclosure to authorities, family members, or others of experiences of gender-based sexual violence may be inhibited because of shame, religious beliefs, gender power imbalances, or fear of retribution (Babalola et al., 2015; Fontes & Plummer, 2010; Nguyen et al., 2021; Ullman et al., 2008). Negative social reactions or disbelief and lack of trust in state agencies can further discourage disclosure (Ahrens C.E., 2007), resulting in low rates of victims receiving assistance (Sumner S. A., 2015).

For example, a Nigerian study of female victims of sexual violence aged 13 to 24 revealed that only one-third of the victims discussed the incident with someone, with friends (38%) and parents (32%) being the most common confidants. Furthermore, only one in seven victims knew where to seek help, and only 4% received actual assistance. Many victims were afraid of the potential consequences of disclosure (30%), while others did not believe that reporting the incident was necessary (35%) or think that it was a significant issue (16%) (Ikuteyijo et al., 2022; Ikuteyijo, 2022; Nguyen et al., 2021). The latter reasons likely stem from a tacit acceptance, and potentially even an endorsement, of violence against women as a social norm (Gilbert et al., 2022).

*The MTV Shuga Naija* program consisted of a TV serial drama that is supported by radio programming and social media and aimed to challenge sexual violence norms, promote gender equality, and encourage attitudinal and behavior change regarding sexual violence among young people. Storylines portrayed the experiences of young women who were victims of sexual violence, including how they encountered stigma and sought justice and support from adults and peers. The program showcased how greater dialogue, both publicly and with parents and other adults, could foster lesser tolerance of violence and greater assistance for victims, leading to broader social change.

The advantages of the entertainment-education format are numerous. Such media initiatives have low access costs, enabling wide reach with audiences through television, radio, and film. Compelling storylines can make content relatable and engender emotional connections with characters and situations, as well as present positive role models whose attitudes and behaviors can be emulated by viewers and listeners. Captivating narratives can stimulate conversations about difficult topics within communities and social networks, contributing to social change (Singhal, 1999, 2004; Singhal & Rogers, 2001; Usdin, 2004; Yue Z, 2019).

As evidence of the effectiveness of *MTV Shuga*’s entertainment education approach, an earlier evaluation of prior seasons, which focused on HIV/AIDS and safer sex, used a randomized controlled trial to demonstrate positive causal impacts of MTV Shuga programs on attitudes towards domestic violence, as well as on HIV/AIDS knowledge and attitudes, risky sexual behavior, and reductions in STDs among female youth (Banerjee, 2019). A 2018 study linked exposure to Season 3 of *MTV Shuga,* which contained a subplot about a married couple with a violent husband, with improved attitudes among male viewers, who were 21 percent less likely to justify domestic violence (Banerjee, 2019).

This research seeks to further explore the impacts of *MTV Shuga*, focusing specifically on issues of sexual violence among Nigerian youth. To further contribute to the evidence base for entertainment education programming, the study uses a quasi-experimental approach that compares outcomes for youth in Lagos, where programming with themes of sexual violence aired, with youth in the states of Kaduna and Kano, where such programming was absent. The principal research objective is to estimate whether *MTV Shuga Naija* was effective in reducing victim-blaming attitudes, increasing reporting of experiences of sexual harassment and sexual violence and increasing interpersonal discussion about sexual violence among Nigerian youth and their parents. We use an analytical approach, doubly robust, fixed effects Difference-in-Differences (DID) models, that adjusts for possible confounding due to unobserved heterogeneity between treatment and comparison individuals, which could otherwise potentially lead to over- or under-estimates of the true effectiveness of *MTV Shuga Naija*.

## Methods

### Intervention

The design and content of *MTV Shuga Naija* is guided by the Theory of Planned Behavior (Ajzen, 1991), which posits that changes in the precursors to targeted behaviors – attitudes, subjective norms, and perceived behavioral control – lead eventually to changes in behavioral intention and, in turn, to changes in behaviors *Shuga* programming is designed to include content specific to behavioral beliefs, normative beliefs, and control beliefs.

The *MTV Shuga Naija* television and radio dramas confront challenging issues by featuring diverse characters who express a variety of attitudes, norms, and behaviors. The show presents new, positive attitudes, norms, and behaviors for young people to emulate. At the same time, it also depicts traditional or harmful attitudes, norms, and behaviors, demonstrating how these can negatively affect youth. On the issue of sexual violence, storylines illustrated how behaviors such as respecting the autonomy of sexual partners, supporting victims of sexual violence, and not engaging in sexual violence led to better outcomes, while also depicting the possible negative consequences of harmful behaviors, both for victims and for perpetrators of sexual violence.

In the 2018-2020 seasons of *MTV Shuga Naija*, issues of sexual violence were portrayed in two key storylines – the rape of a high school girl at a party thrown by older boys and the rape of a sexually experienced “video vixen” and aspiring singer by an influential music producer. Smaller storylines – such as a teacher soliciting sexual favors from a female student to improve her grades or a male student suggestively touching female students – depicted forms of sexual harassment. Storylines showed the victims struggling with the after-effects of sexual assault and harassment, modeling proactive behaviors to seek justice, and ultimately prevailing over entrenched norms and social structures, thereby offering viewers a path to deal with their own experiences of sexual violence.

The target population for *MTV Shuga Naija* was youth in the states of Kaduna, Kano, and Lagos, although the content in Lagos was broader than in the northern two states. In Lagos, storylines touched not just on sexual harassment and violence but also on sexual education and reproductive health. From March-April 2018, ten 22-minute episodes of the television serial drama were aired on MTV satellite and NTA2 Lagos (Nigeria Television Authority) and again 18 months later. The episodes on NTA2 were available only for viewers in the Lagos area while those airing on MTV satellite were potentially viewable anywhere in the country, although access was generally considerably lower outside of Lagos. Episodes also aired on the www.mtvshuga.com website and on the MTV Shuga YouTube channel. The episodes were complemented by a radio drama, social media (e.g., mtvshuga.com website, Facebook, Instagram, Twitter), and peer education sessions with youth in Lagos. A second 8-episode television drama series that continued the storylines in the earlier series were shown in October/November 2019. This series aired on MTV satellite, MTV Base, Startimes TV, Pop Central, NTA, ONTV, and EBONYLIFE TV, alongside a more extensive radio drama, peer education, and social media.

Beginning in 2019, *MTV Shuga* commenced limited program activities in Kaduna and Kano, focused solely on reproductive health and absent content and storylines about sexual violence. Activities included a television magazine, a radio drama, social media, and peer education. Themes of this programming

### Data

To assess the effects of *Shuga* on the target groups, surveys were conducted among a randomly sampled cohort of urban youth. Consistent with the stepwise introduction of the intervention, youth were interviewed at three points in time: (1) prior to the implementation of *Sahnuyga Naija* activities in any of the states (baseline), (2) after implementation of *Shuga Naija* activities in Lagos but not elsewhere (midline), and (3) after the implementation of *Shuga Naija* activities in all three states. We focus only on the baseline and endline waves of data because at midline, many of the critical story arcs regarding sexual violence were still emerging, meaning that key learnings for youth were not fully revealed.

At baseline, the target sample size for the study was computed as 2,000 females and 1,000 males between the ages of 14-24 years. With permission from the Bill and Melinda Gates Institute of the Johns Hopkins University, Baltimore, the Center for Research, Evaluation Resources, and Development (CRERD) used the Performance Monitoring for Accountability 2020 (PMA2020 2017) Nigeria sample frame in Kano, Lagos, and Kaduna states to select urban clusters for the *Shuga* survey. Utilization of PMA2020 enumeration areas (EAs), which were randomly selected from each state’s census enumeration areas, meant that there was a previously established and recent sampling frame of all households located in PMA2020 EAs.

At baseline, in-person interviews were completed with 1,900 females and 986 males. As with most panel surveys, attrition of respondents occurred across survey waves. Of the baseline participants, 70% agreed to be re-contacted for subsequent interviews, and attempts were made to re-contact these individuals for the endline survey.

Because of contact restrictions due to the COVID-19 pandemic and to comply with COVID-19 regulations, data collection for the endline survey was undertaken through phone interviews. Phone calls were made to all respondents in the baseline and midline database who had valid phone numbers and consented to callbacks.^1^ The switch to phone surveys compounded earlier attrition, resulting in an endline sample of 899 respondents. Of these, only 641 in total (399 females and 242 males), participated in all three waves.

### Survey Instruments

The *MTV Shuga Naija* survey instruments were designed to capture information on *Shuga* exposure and indicators from the Theory of Planned Behavior (Ajzen 1991) that guided the development of *MTV Shuga Naija* programming. The questionnaire focused on issues that were specifically addressed by the storylines for each of the *MTV Shuga Naija’s* main characters. For example, questions were asked about: 1) behaviors or experiences (e.g., “Has someone ever touched you in a way that made you feel uncomfortable?” “Have you ever talked with your parents or family members about sexual violence?”); 2) attitudes (e.g., “It’s not rape if a girl doesn’t fight back”); 3) subjective norms (e.g., “Most of my friends think I should use condoms when having sexual intercourse”); 4) perceived behavioral control (e.g., “How easy or difficult would it be for you to use condoms with a sexual partner?”); and 5) behavioral intentions (e.g., “During the next 3 months, how likely is it that you will try to persuade your partner to use condoms every time you have sex?”).

Because the endline survey was conducted by phone, a shorter questionnaire was required to minimize respondent fatigue and drop-off. Questions focused on a smaller set of indicators from the TPB that were deemed essential to cover the theory’s broad domains and behavioral outcomes (e.g., contraceptive use, HIV testing, experiences and discussions of sexual violence).

### Human Subjects Research Approval

For the baseline survey, National Health Research Committee (NHREC) approval was obtained on January 24, 2018 with NHREC/01/01/2007-24/01/2018. Institutional Review Board (IRB) approval from the Tulane University Biomedical IRB was granted on February 18, 2018. For the midline survey, IRB approval from the NHREC was obtained, which spans August 23, 2019 through August 22, 2020 (NHREC/01/01/2007-23/08/2019B). Approval from the Tulane University Biomedical IRB was granted on September 20, 2019. For the endline survey, CRERD received approval on July 9, 2020. Approval from the Tulane University Biomedical IRB was granted on August 5, 2020.

### Statistical Analysis

Our analytical approach is to estimate two-period, two-group difference-in-differences models, comparing changes over time in key outcomes in the treatment area of Lagos relative to those in the comparison areas of Kaduna and Kano. The primary assumption underlying the DID model is that, in the absence of the treatment (*MTV Shuga Naija*), average outcomes in treatment and control areas would have followed the same trend from baseline to endline. Consequently, any differences observed in those trends at endline are attributed to the causal influence of *MTV Shuga Naija*. In the DID models, we include a time dummy variable (baseline wave=0, endline wave=1), a binary variable indicating if a person resides in the treatment area of Lagos versus the control areas of Kaduna and Kano, and an interaction between the two. The effect of the interaction terms is the Average Treatment Effect on the Treated (ATT). More explicitly, this is a measure of the mean impact of *MTV Shuga Naija* on the population in areas that were exposed to it.

We make use of the panel nature of the data set to estimate a fixed effects regression model for each outcome related to experience of sexual violence and for attitudes towards victims of sexual violence. These models capture differences between treatment and control groups through the group fixed effect and control for any temporal factors that affect both treatment and control groups equally through the time fixed effect. Because these models also contain an individual fixed effect, any time invariant variable for an individual drops out of the model. This means that the binary variable for living in the treatment or comparison area drops out, although the interaction with the binary survey wave variable remains.

Because we are concerned about the validity of the parallel trends assumption, we use a method that combines the fixed-effects model with inverse probability weighting (IPW). This approach adjusts for potential confounding by weighting observations by the inverse of their probability of receiving the treatment based on their surveyed characteristics (Joffe et al., 2004). The resulting estimator, known as the doubly robust (DR) estimator, (Callaway, 2021; Lunceford & Davidian, 2004) is robust to misspecification of either the outcome (fixed effects) regression model or the IPW model. The DR model in effect produces a weighted combination of the estimates from the outcome and IPW models, with the weights reflecting the degree of overlap between the treatment and control groups. The DR estimator consistently estimates the treatment effect parameters provided either the regression or the propensity score is correctly specified.

While the dependent variables in each case are binary, we estimate the models using a linear fixed effects regression model in order to avoid the incidental parameters problem inherent in non-linear fixed effects models (Chamberlain, 1980). This problem arises because nonlinear fixed effects models treat individual fixed effects as parameters to be estimated, and, when the number of time periods is small, these models may produce inconsistent estimates of the individual fixed effects, in turn leading to inconsistent estimates of other parameters in the model. Consequently, estimation of the ATT using standard statistical software like Stata is not straightforward,

By using the linear fixed effects model, we are able to directly identify the ATT, the primary focus of our study, as the coefficient on the interaction term for treatment area and survey wave. However, we are unable to estimate average predicted probabilities for observed values of covariates (e.g., the predicted outcome for those receiving the treatment versus those not receiving the treatment) because the individual-specific effect, the intercept for each individual across all time periods, is differenced out of the model through the within transformation or “demeaning” of the data inherent to fixed effects models. Once these individual-specific effects are differenced out in the fixed effects models, we can ascertain changes in the outcome variable from changes in individual covariates but not predictions of the outcome variable for different values of covariates (Wooldridge, 2002). Consequently, we present only the average treatment effects from the DID models.

All models were estimated in Stata 18.0 using the *drdid* command developed by Sant’Anna (2020). This estimator implements the doubly robust difference-in-differences estimator for the average treatment effect with the inverse probability of tilting (IPT) and weighted least squares (WLS).

We also sought to address the possible influence of changes in the sampling procedure associated with moving from face-to-face interviews, as was done for the baseline wave, to phone interviews, as was necessitated by COVID-19 in the endline wave. Our strategy involves adjusting our sample weights through an iterative proportional fitting procedure (Bergmann, 2011). Because the endline sample was restricted to respondents who had consented to being re-contacted by phone, it contained respondents with a different socioeconomic profile on average than the samples for the first two waves. The unweighted endline sample, for example, was found to be more educated on average; 27.7% of females and 34.5% of males in the endline had post-secondary education as compared with 11.8% and 12.5% respectively at midline.

Using the iterative proportional fitting procedure (also known as “survey raking”), we adjusted our sample weights for endline respondents so that the demographic profile matched that of the baseline sample in terms of gender, state, education and marital status. Sample weights for the endline sample were iteratively adjusted to minimize the discrepancies with the known population distribution of characteristics of the baseline sample until the differences between the two samples converged to pre-specified tolerance level.^2^ Survey raking was conducted using the *ipfraking* and *ipfweight* commands in Stata 17 (Kolenikov, 2014, 2019).

## Results

### Descriptive Results

The characteristics of the sample changed from baseline to endline, with most of the changes attributable to maturation and transitions to adulthood. All respondents aged by three years from baseline to endline; the share of the female sample that was aged 13-17 years decreased from 51% at baseline to 25% at endline in the treatment area (Lagos) and from 42% to 18% in the comparison area (Kaduna and Kano). Similar patterns were observed in the male sample.

The female sample in the treatment areas had higher educational attainment at both baseline and endline relative to the comparison sample. At both baseline and endline, females in Lagos were approximately 14 percentage points more likely to have completed secondary education relative to females in the comparison areas (Treatment: 85% at baseline, 81% at endline; Comparison: 72% at baseline, 67% at endline). For males, educational attainment was higher in the treatment area at baseline (75% of males with secondary education in the treatment area versus 59% in the comparison area) but by endline greater parity was evident (79% of males with secondary education in the treatment area versus 82% in the comparison areas).

Across all areas, few males, at both baseline and endline, reported that they were currently married, consistent with patterns of considerable later ages at marriage for males relative to females in Nigeria. Females in the comparison areas were approximately three times more likely to be married at endline than females in the treatment area. Relatedly, roughly one in four women in the comparison areas had a child by the endline versus only 8% of women in the treatment area. Less than 3% of men in the comparison areas reported having a child at endline, relative to 75 of men in the treatment areas.

**Table 1a.** Sample Characteristics, by Survey Wave and Sex (with raked weights) Females.

|  | Comparison/North |  | Treatment/Lagos |  |
| --- | --- | --- | --- | --- |
|  | Baseline<br>% | Endline<br>% | Baseline<br>% | Endline<br>% |
| **Age at last birthday** |  |  |  |  |
| 13-17 | 42.2 | 17.5 | 51.3 | 24.7 |
| 18-21 | 36.7 | 48.0 | 24.6 | 44.1 |
| 22+ | 21.1 | 34.4 | 24.1 | 31.2 |
| **Level of Education** |  |  |  |  |
| None | 6.3 | 5.6 | 0.0 | 4.7 |
| Primary | 9.5 | 18.3 | 0.7 | 0.8 |
| Secondary | 71.9 | 67.4 | 85.4 | 80.7 |
| Higher | 12.3 | 8.7 | 13.9 | 13.8 |
| **Marital status** |  |  |  |  |
| Currently married/living with someone | 31.4 | 25.5 | 8.3 | 7.5 |
| Not married | 68.6 | 74.5 | 91.7 | 92.5 |
| **Age at first sex** |  |  |  |  |
| Never sex | 64.4 | 62.0 | 61.6 | 56.4 |
| Don't Know/Refused | 0.0 | 3.7 | 1.9 | 2.7 |
| Younger than 15 | 8.2 | 2.8 | 3.2 | 0 |
| 15-17 | 12.5 | 14.4 | 9.1 | 6.3 |
| 18-21 | 13.2 | 14.8 | 21.7 | 30.7 |
| 21+ | 1.7 | 2.4 | 2.4 | 4.0 |
| **Number of children ever born** |  |  |  |  |
| 0 | 73.2 | 75.6 | 90.1 | 92.4 |
| 1 | 6.0 | 6.0 | 6.9 | 5.5 |
| 2 | 8.5 | 6.5 | 2.1 | 1.9 |
| 3+ | 12.3 | 12.0 | 0.8 | 0.3 |
| Own a phone? | 51.4 | 79.8 | 62.6 | 84.0 |
| Ever watch TV? | 79.1 | 90.1 | 97.0 | 97.8 |
| Ever listen to the radio? | 67.3 | 58.4 | 57.7 | 59.4 |
| N | 386 | 390 | 183 | 184 |

**Table 1b.** Sample Characteristics, by Survey Wave and Sex (with raked weights), Males.

|  | Comparison/North |  | Treatment/Lagos |  |
| --- | --- | --- | --- | --- |
|  | Baseline | Endline | Baseline | Endline |
|  | % | % | % | % |
| **Age at last birthday** |  |  |  |  |
| 13-17 | 32.5 | 8.9 | 46.6 | 27.0 |
| 18-21 | 36.6 | 42.7 | 25.9 | 37.1 |
| 22+ | 30.9 | 48.5 | 27.5 | 35.9 |
| **Level of Education** |  |  |  |  |
| None | 0.4 | 0.0 | 3.4 | 0.0 |
| Primary | 20.7 | 5.3 | 3.1 | 0.0 |
| Secondary | 58.9 | 81.8 | 75.4 | 78.8 |
| Higher | 20.0 | 13.0 | 18.1 | 21.2 |
| **Marital status** |  |  |  |  |
| Currently married/living with someone | 0.8 | 4.2 | 4.8 | 1.7 |
| Not married | 99.2 | 95.8 | 95.2 | 98.3 |
| **Age at first sex** |  |  |  |  |
| Never Sex | 86.6 | 73.2 | 63.9 | 47.1 |
| Don't Know/Refused | 1.2 | 6.7 | 1.5 | 3 |
| Younger than 15 | 1.7 | 0.5 | 4.4 | 3.4 |
| 15-17 | 3.8 | 2.2 | 10.6 | 18.5 |
| 18-21 | 6.1 | 13.4 | 17.9 | 23.6 |
| 21+ | 0.5 | 4.0 | 1.8 | 4.4 |
| **Number of children ever born** |  |  |  |  |
| 0 | 99.4 | 97.5 | 94.5 | 93.2 |
| 1 | 0.0 | 1.0 | 2.7 | 1.9 |
| 2 | 0.0 | 0.9 | 0.7 | 1.6 |
| 3+ | 0.6 | 0.7 | 2.1 | 3.4 |
| Own a phone? | 73.6 | 83.1 | 70.0 | 88.1 |
| Ever watch TV? | 89.6 | 94.5 | 99.5 | 96.4 |
| Ever listen to the radio? | 67.0 | 62.6 | 71.1 | 80.3 |
| N | 386 | 390 | 183 | 184 |

At both baseline and endline, the majority of youth owned a cellphone. Women in the treatment area appear to have gotten a cellphone sooner than women in the comparison areas; 63% of women in Lagos owned a cellphone at baseline versus 51% of women in the comparison areas. By endline, roughly eight in ten women in both areas (treatment: 84%, comparison: 80%) owned a cellphone. Approximately equal percentages of males in treatment and comparison areas owned cellphones at both baseline and at endline.

The majority of youth report exposure to both radio and television, with television being the more commonly used than radio. At endline, women in treatment areas were slightly more likely to watch television than women in comparison areas (treatment: 98%, comparison: 90%) but equally likely to listen to the radio (treatment: 59%, comparison: 58%). Males were equally likely to watch television at endline (treatment: 96%, comparison: 95%), but males in the treatment area were more likely to listen to the radio (treatment: 80%, comparison: 63%).

### Sexual Violence

From baseline to endline, there were significant changes in experiences of sexual harassment, attitudes towards victims of sexual violence, and the likelihood of discussing sexual violence with family members for both females and males (Tables 2a and 2b).

**Table 2a.**
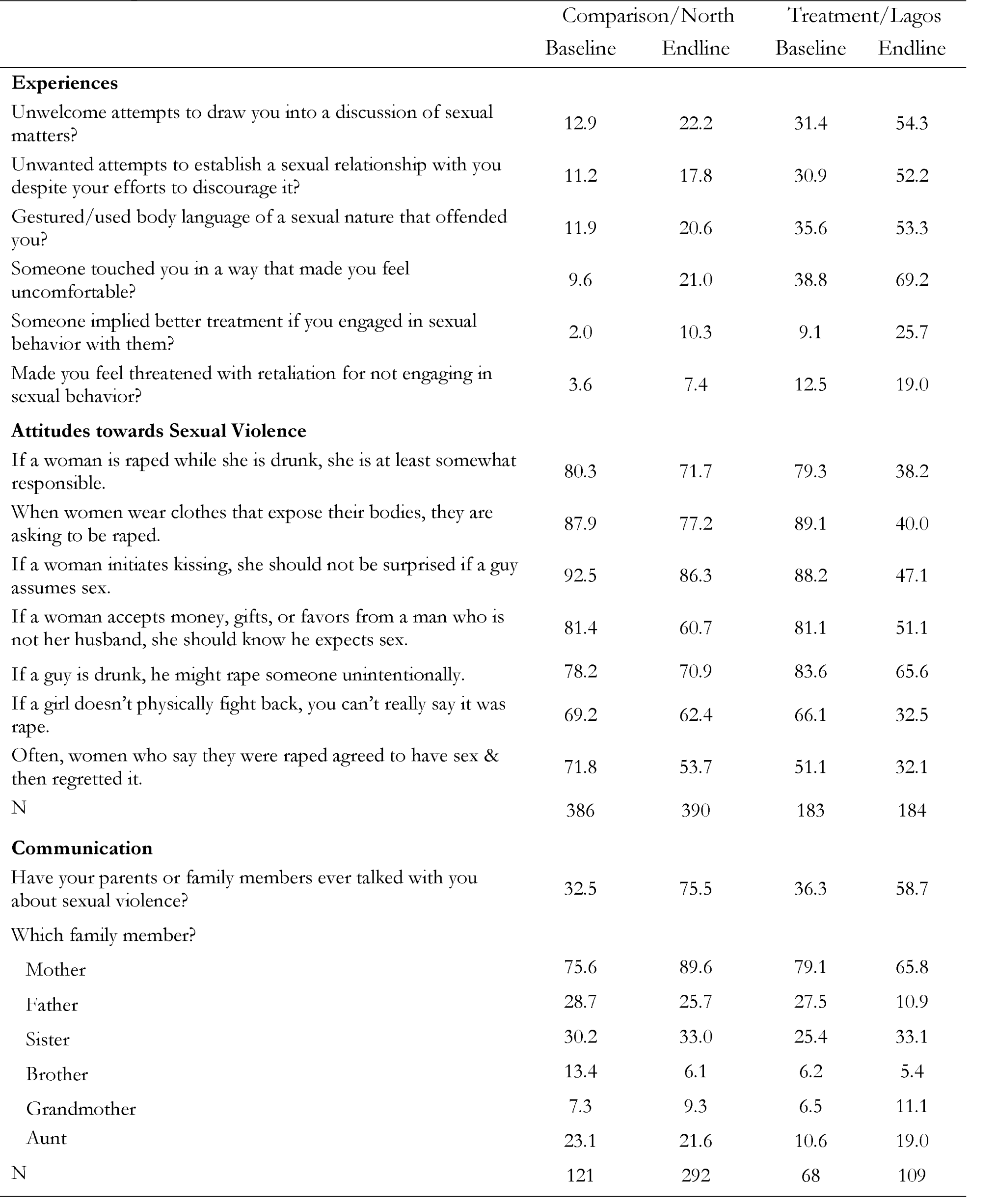
Experiences of sexual harassment/violence and attitudes towards sexual violence, Females.

|  | Comparison/North |  | Treatment/Lagos |  |
| --- | --- | --- | --- | --- |
|  | Baseline | Endline | Baseline | Endline |
| **Experiences** |  |  |  |  |
| Unwelcome attempts to draw you into a discussion of sexual matters? | 12.9 | 22.2 | 31.4 | 54.3 |
| Unwanted attempts to establish a sexual relationship with you despite your efforts to discourage it? | 11.2 | 17.8 | 30.9 | 52.2 |
| Gestured/used body language of a sexual nature that offended you? | 11.9 | 20.6 | 35.6 | 53.3 |
| Someone touched you in a way that made you feel uncomfortable? | 9.6 | 21.0 | 38.8 | 69.2 |
| Someone implied better treatment if you engaged in sexual behavior with them? | 2.0 | 10.3 | 9.1 | 25.7 |
| Made you feel threatened with retaliation for not engaging in sexual behavior? | 3.6 | 7.4 | 12.5 | 19.0 |
| **Attitudes towards Sexual Violence** |  |  |  |  |
| If a woman is raped while she is drunk, she is at least somewhat responsible. | 80.3 | 71.7 | 79.3 | 38.2 |
| When women wear clothes that expose their bodies, they are asking to be raped. | 87.9 | 77.2 | 89.1 | 40.0 |
| If a woman initiates kissing, she should not be surprised if a guy assumes sex. | 92.5 | 86.3 | 88.2 | 47.1 |
| If a woman accepts money, gifts, or favors from a man who is not her husband, she should know he expects sex. | 81.4 | 60.7 | 81.1 | 51.1 |
| If a guy is drunk, he might rape someone unintentionally. | 78.2 | 70.9 | 83.6 | 65.6 |
| If a girl doesn't physically fight back, you can't really say it was rape. | 69.2 | 62.4 | 66.1 | 32.5 |
| Often, women who say they were raped agreed to have sex & then regretted it. | 71.8 | 53.7 | 51.1 | 32.1 |
| N | 386 | 390 | 183 | 184 |
| **Communication** |  |  |  |  |
| Have your parents or family members ever talked with you about sexual violence? | 32.5 | 75.5 | 36.3 | 58.7 |
| Which family member? |  |  |  |  |
| Mother | 75.6 | 89.6 | 79.1 | 65.8 |
| Father | 28.7 | 25.7 | 27.5 | 10.9 |
| Sister | 30.2 | 33.0 | 25.4 | 33.1 |
| Brother | 13.4 | 6.1 | 6.2 | 5.4 |
| Grandmother | 7.3 | 9.3 | 6.5 | 11.1 |
| Aunt | 23.1 | 21.6 | 10.6 | 19.0 |
| N | 121 | 292 | 68 | 109 |

**Table 2b.** Experiences of sexual harassment/violence and attitudes towards sexual violence, Males.

|  | Comparison/North |  | Treatment/Lagos |  |
| --- | --- | --- | --- | --- |
|  | Baseline | Endline | Baseline | Endline |
| **Experiences** |  |  |  |  |
| Unwelcome attempts to draw you into a discussion of sexual matters? | 17.6 | 33.8 | 21.5 | 53.8 |
| Unwanted attempts to establish a sexual relationship with you despite your efforts to discourage it? | 20.3 | 40.2 | 20.0 | 51.2 |
| Gestured/used body language of a sexual nature that offended you? | 26.3 | 35.8 | 21.6 | 47.3 |
| Someone touched you in a way that made you feel uncomfortable? | 23.8 | 48.8 | 30.6 | 65.1 |
| Someone implied better treatment if you engaged in sexual behavior with them? | 4.6 | 19.7 | 10.4 | 20.3 |
| Made you feel threatened with retaliation for not engaging in sexual behavior? | 7.7 | 22.9 | 10.2 | 14.9 |
| **Attitudes towards Sexual Violence** |  |  |  |  |
| If a woman is raped while she is drunk, she is at least somewhat responsible. | 83.4 | 68.4 | 85.0 | 56.8 |
| When women wear clothes that expose their bodies, they are asking to be raped. | 89.0 | 89.8 | 91.3 | 61.0 |
| If a woman initiates kissing, she should not be surprised if a guy assumes sex. | 92.4 | 78.8 | 95.5 | 80.9 |
| If a woman accepts money, gifts, or favors from a man who is not her husband, she should know he expects sex. | 83.0 | 67.0 | 87.1 | 68.2 |
| If a guy is drunk, he might rape someone unintentionally. | 70.6 | 63.5 | 67.9 | 71.0 |
| If a girl doesn't physically fight back, you can't really say it was rape. | 78.6 | 66.3 | 75.6 | 52.9 |
| Often, women who say they were raped agreed to have sex & then regretted it. | 60.7 | 62.2 | 57.9 | 60.6 |
| N | 222 | 222 | 95 | 95 |
| **Communication** |  |  |  |  |
| Have your parents or family members ever talked with you about sexual violence? | 8.0 | 33.5 | 20.8 | 50.2 |
| Which family member? |  |  |  |  |
| Mother | 29.6 | 59.6 | 59.9 | 54.0 |
| Father | 7.4 | 53.2 | 66.7 | 43.0 |
| Sister | 31.3 | 20.8 | 14.1 | 9.7 |
| Brother | 48.7 | 44.5 | 26.4 | 19.9 |
| Grandmother | 0.0 | 0.2 | 1.3 | 0.0 |
| Aunt | 0.0 | 12.4 | 1.3 | 5.1 |
| N | 17 | 81 | 20 | 49 |

Reporting of experiences of sexual harassment increased across all categories for both females and males, potentially reflecting a growing awareness of the issue. Reporting of unwelcome attempts to draw participants into a discussion of sexual matters increased from 16.1% to 33.0% for females and 16.3% to 39.7% for males. Unwanted attempts to establish a sexual relationship also increased from 13.7% to 29.4% for females and 17.2% to 43.7% for males. Touching in a way that made participants feel uncomfortable rose from 14.0% to 37.5% for females and 22.7% to 53.7% for males.

Attitudes towards victims of sexual violence generally improved from baseline to endline. The belief that a woman is at least somewhat responsible for being raped while drunk decreased from 80.5% to 60.3% for females and 82.3% to 64.6% for males. The view that women are asking to be raped when wearing revealing clothing dropped from 86.3% to 64.7% for females and decreased from 86.7% to 81.4% for males. Even with these shifts, there remains a significant gender disparity in some beliefs. For example, the belief that women wearing revealing clothing are asking to be raped remains high among males.

The likelihood of discussing sexual violence with family members increased dramatically. Reporting of conversations with family about sexual violence rose from 31.2% to 70.0% for females and 9.0% to 38.5% for males. Mothers were the most common family members to engage in these discussions, with the percentage increasing from 73.7% to 82.9% for females and 42.1% to 56.4% for males. Discussions with fathers also increased, from 23.6% to 21.3% for females and 25.1% to 49.2% for males. These results coincide with a key *MTV Shuga Naija* theme: engaging parents, family members, and society at large in open conversations about sexual violence may contribute to improved awareness and understanding and ultimately help reduce the occurrence of sexual harassment and assault.

### Difference-in-Differences Models

Overall, the multivariate DID results provide support for the effects of MTV Shuga Naija on attitudes towards sexual violence and disclosure of experiences of sexual harassment and sexual violence, but lesser support for increased communication with family members about sexual violence.

The program had dramatic and demonstrable effects on shifting harmful attitudes about sexual violence and victims, particularly among females, with notable decreases in victim-blaming attitudes in instances of rape under various scenarios, such as when a woman is drunk [ATT=-32.0pp, 95% CI: -43.4pp, -20.7pp], initiates kissing [ATT=-29.3pp, 95% CI: -38.9pp, -19.6pp], or does not fight back [ATT=-28.3pp, 95% CI: -40.2pp, -16.4pp], and attitudes justifying rape such as “when women where clothes that expose their bodies, they are asking to be raped” [ATT=-36.6pp, 95% CI: - 46.8pp, -26.3pp] or “if a guy is drunk, he can rape someone unintentionally” [ATT=-12.0pp, 95% CI: -23.0pp, -1.1pp]. The effects for males were less pronounced but still statistically significant for justification for rape when a woman is wearing revealing clothing [ATT=-31.8pp, 95% CI: -45.6pp, - 18.1pp] and for characterization of rape as requiring that a girl fight back [ATT=-19.3pp, 95% CI: - 35.6pp, -3.0pp]. These latter results may indicate that male attitudes are harder to shift or that different types of messages, sources of messages, or other interventions may be more effective with males.

In Lagos, both males and females reported a significant increase in disclosure to interviewers of experiences of sexual harassment, such as unwelcome attempts to discuss sex and offensive gesturing, relative to youth in the comparison states of Kaduna and Kano. For females, significant effects were observed for “unwanted attempts at a sexual relationship” [ATT=12.3pp, 95% CI: 1.1pp, 23.6pp], and “touched you in an uncomfortable way” [ATT=14.8pp, 95% CI: 3.2pp, 26.5pp]. For males, statistically significant effects were observed for “unwanted attempts at a sexual relationship” [ATT=18.5pp, 95% CI: 0.5pp, 36.4pp] and “gestured in an offensive way” [ATT=24.7pp, 95% CI: 8.1pp, 41.4pp]. These results suggest improved awareness and recognition of sexual harassment due to the program, leading to increased reporting.

A key objective of*MTV Shuga Naija* was to remove the stigma around victims of sexual violence and to promote open dialogue so that attitudes could shift, norms surrounding appropriate sexual contact could be improved, and victims would be more willing to seek services post-experience. While dialogue with family members about sexual violence indeed improved in the treatment areas, the DID models found that improvements in communication in the comparison areas actually outpaced those in the treatment areas among females [ATT=-19.9pp, 95% CI: -32.1pp, -7.8pp]. Interpretation of this finding is challenging but may lie in the phrasing of the question (“Have your parents or family members ever talked with you about sexual violence?), which provides little information on who initiated the conversation or what the content of the conversation was. It is possible that such conversations might even have perpetuated or articulated some of the same victim-blaming stereotypes and attitudes as admonishments and warnings toward youth.

**Table 3a.**
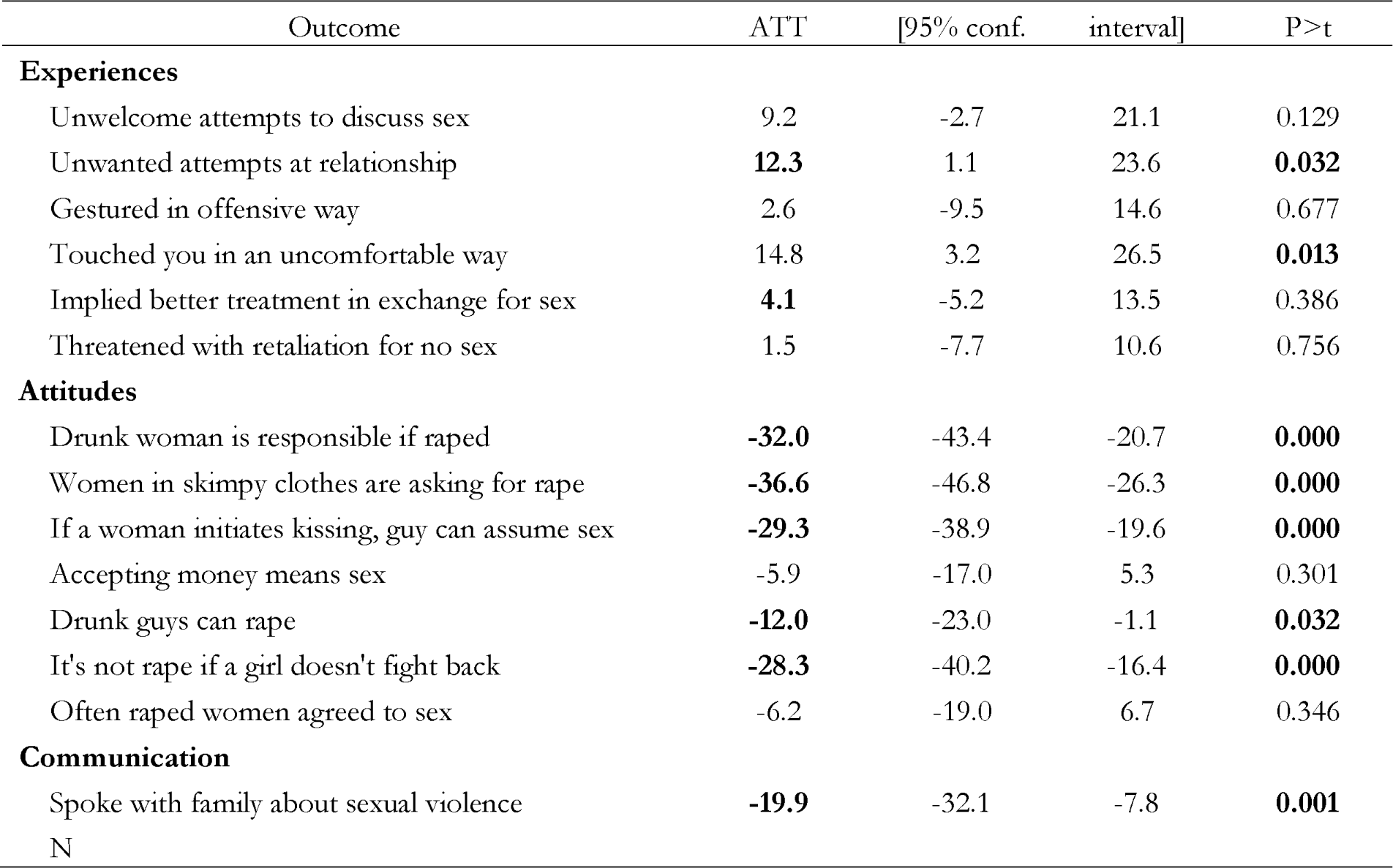
Doubly-Robust Inverse Probability Weighted (Balanced Panel) Regression Models, Females.

**Table 3a.** Doubly-Robust Inverse Probability Weighted (Balanced Panel) Regression Models, Males.

| Outcome | ATT | [95% conf. interval] | P>t |
| --- | --- | --- | --- |
| **Experiences** |  |  |  |
| Unwelcome attempts to discuss sex | 18.4 | -2.1 38.9 | 0.078 |
| Unwanted attempts at relationship | **18.5** | 0.5 36.4 | **0.044** |
| Gestured in offensive way | **24.7** | 8.1 41.4 | **0.004** |
| Touched you in an uncomfortable way | 13.3 | -4.2 30.7 | 0.137 |
| Implied better treatment in exchange for sex | 5.5 | -7.3 18.3 | 0.402 |
| Threatened with retaliation for no sex | -10.9 | -26.2 4.3 | 0.160 |
| **Attitudes** |  |  |  |
| Drunk woman is responsible if raped | **-25.3** | -42.1 -8.6 | **0.003** |
| Women in skimpy clothes are asking for rape | **-31.8** | -45.6 -18.1 | **0.000** |
| If a woman initiates kissing, guy can assume sex | -1.3 | -12.6 10.1 | 0.827 |
| Accepting money means sex | 3.0 | -12.9 19.0 | 0.708 |
| Drunk guys can rape | -2.7 | -21.7 16.4 | 0.785 |
| It's not rape if a girl doesn't fight back | **-19.3** | -35.6 -3.0 | **0.020** |
| Often raped women agreed to sex | 4.4 | -13.9 22.8 | 0.637 |
| **Communication** |  |  |  |
| Spoke with family about sexual violence | 2.2 | -15.3 19.7 | 0.806 |
| N |  |  |  |

## Discussion

The compelling and dramatic storylines represented in the MTV *Shuga Naija* serial drama, coupled with numerous opportunities for multi-way dialogue through social and other media, are targeted to youth in Nigeria, where women frequently have limited autonomy and experiences of sexual harassment and sexual violence are commonplace. *MTV Shuga Naija* aims to shift norms that are accepting of such violence, to fostearttitudes thatare supportive of victims, and to empower youth to speak up and seek assistance for incidents of violence. *MTV Shuga Naija* was implemented from 2018-2020, which was an extremely opportune time given that the Me Too movement in Nigeria was also getting considerable attention in the press. The results presented here contribute to the scant evidence base surrounding the use of entertainment education to improve sexual violence attitudes, norms, and disclosure.

This evaluation is unique in that it demonstrates a link between multichannel social and behavior change programming and attitudes towards sexual violence. To date, there has been limited evidence of such a link in contexts similar to the areas under study here (Bott et al., 2005). The WHO and PAHO have highlighted the important role that mass media entertainment education strategies can have in influencing gender norms, community responses, and individual attitudes towards intimate partner violence (World Health Organization & Pan American Health Organization, 2012).

The findings indicating that *MTV Shuga Naija* has been successful in shifting norms and attitudes are indeed encouraging, and yet there appears to still be quite a long way to go. Even among those exposed to the campaign, there was still a high prevalence of victim blaming attitudes. Nearly eight of 10 youth exposed to *MTV Shuga Naija* agreed that, if a woman initiates kissing, a guy can assume that sex will occur, and more than half of the youth respondents believed that it is not rape if a woman does not fight back. More progress is needed in order for victims of sexual violence to feel confident of support and understanding, and ideally for victimization to become less common.

The approaches taken in Shuga’s programmatic design and message design are important. The overall programmatic design was based in entertainment-education change (Singhal, 1999, 2004; Singhal & Rogers, 2001; Usdin, 2004; Yue Z, 2019). The observed beneficial effects of Shuga in this study can be theorized to function through the mechanisms that permit entertainment-education to grab the attention of and captivate audience members, while also infusing in them norms and information pertinent to gender-based violence. Moreover, the message design was based in the Theory of Planned Behavior (Ajzen, 1991). The television and radio dramas purposefully included content specific to behavioral beliefs, normative beliefs, and control beliefs, which are conceived to be core processes, respectively, in the development of attitudes, subjective norms, and perceived behavioral control. Whereas this study did not implement measures of each of these steps in the context of gender-based violence, the evaluation provides evidence generally in line with the expected outcomes of this theory and underscores the importance of using programmatic and message-design theory in the development and evaluation of health campaigns.

This research points to areas where additional investigation is warranted, including research that focuses specifically on understanding the factors that contribute to the persistence of victim-blaming attitudes and misconceptions about consent. Further, there remains a paucity of studies that seek to understand the challenges of reporting incidences of sexual violence to security outfits including police. We suggest research that explores the role of cultural, societal, and familial influences in shaping these attitudes and identifying ways to address these factors more effectively through targeted interventions. In addition, future studies of entertainment-education could investigate the impact of different message framing, content, and delivery formats in changing attitudes towards victim-blaming and consent and identifying best practices for developing more persuasive campaigns. The Theory of Planned Behavior provides one widely used framework for message design, but future entertainment-education research in this area could employ others, as well. Additionally, studies could evaluate the effectiveness of integrating other strategies, such as community-based initiatives, educational programs, and policy changes, to reinforce the messages conveyed through entertainment-education programs like *MTV Shuga Naija*.

A longer-term perspective is also warranted. The *MTV Shuga Naija* campaign achieved some impressive results in the few years that it aired. Important empirical issues remain, including assessing the long-term effects of such campaigns on behavior change and exploring whether the impact of exposure to these campaigns persists over time or diminishes without continued reinforcement.

### Limitations and Future Directions

It is important to note that this evaluation faced several challenges. In addition to selective attrition and the switch from face-to-face interviews to phone interviews, there may also have been issues related to differences in the cultures and values between the treatment and comparison areas. The populations of the northern states of Kaduna and Kano are predominantly Muslim, reflected in much lesser autonomy for females,aptterns of early marriage and early child-bearing, and lesser emphasis on schooling and work outside the home for females, in strong contrast to the more western-focused, cosmopolitan populations of the south, including Lagos. The original study design articulated the provision of identical content across all areas, albeit with staggered implementation between the south and the north. However, local governments in the north objected to the explicit content related to sex and sexual violence in *MTV Shuga Naija* programming and prohibited the airing of any material except that focused solely on reproductive health within the context of marriage. These differences in the population characteristics for the treatment and comparison groups may have implications for one of the key assumptions of this analysis – that treatment and control groups would have followed parallel trends in sexual violence attitudes in the absence of the program. While the doubly-robust DID estimation strategy sought to ameliorate the possible influence of these differences, it was simply not feasible to collect more than one pre-intervention round of data that would have permitted a more robust assessments of trends prior to the implementation of *Shuga Naija* activities. Hence an explicit test of the parallel trends assumption was simply not possible.

Second, related to attrition, the sample sizes of respondents, particularly of sexually active males, were quite small at endline. The study was originally powered under the assumption of lower rates of attrition, and as a result, the observed differences required for statistical significance for indicators dependent on sexual activity tended to be quite large. Hence, while *Shuga* may have shifted indicators in the desired direction, our final sample size may have been too small in some cases to observe statistically significant shifts (i.e., at a p-value of below .05).

Finally, starting in March 2020 – seven months before the endline survey – Lagos was placed under stringent COVID-19 lockdowns that were not implemented in the northern states of Kaduna and Kano. This potentially affected the ability of youth in Lagos to interact with others. Hence, changes in the rates of sexual harassment and sexual violence may partially reflect changes in mobility circumstances.

### Conclusion

This evaluation provides evidence of *MTV Shuga Naija*’s effects on attitudes and behaviors related to sexual violence, stigmatization of victims of sexual violence, and open dialogue about addressing sexual violence. During the study period, more rapid attitudinal shifts towards sexual violence and victims of sexual violence were apparent among both male and female youth in the treatment group relative to the comparison group. These results highlight that *MTV Shuga Naija* was able to achieve important effects in a relatively short period of time. It is well established that it takes time for campaign viewers to progress from campaign exposure to the development of revised perceptions and, subsequently, from the attitudinal shifts to the development of revised behavioral patterns. By opening and normalizing dialogue about sexual violence among youth, *MTV Shuga Naija* has set the groundwork for subsequent behavior changes that can help reduce the problem of sexual violence.

## Data Availability

All data produced in the present study are available upon reasonable request to the authors.

## ORCID iDs

Paul Hutchinson https://orcid.org/0000-0002-5435-7799

Christopher E. Beaudoin https://orcid.org/0000-0003-0649-3347

Dominique Meekershttps://orcid.org/0000-0002-7281-3217

Elizabeth Omoluabi. https://orcid.org/0000-0003-1754-5384

Akanni Akinyemi https://orcid.org/0000-0001-8652-9359

## Footnotes

1 At baseline, 1,306 females (68.7%) and 712 males (72.2%) consented to be recontacted.

2 Convergence is achieved if the largest relative difference of the weights in two successive iterations (a full cycle over all raking variables) does not exceed a tolerance of 0.000001.

## References

Ahrens C.E., C. R., Ternier-Thames N.K., Wasco S.M., Sefl T. (2007). Deciding whom to tell: Expectations and outcomes of rape survivors’ first disclosure. Pss.ychology of Women Qua,rterly, 31, 38–49.

Ajzen, I. (1991). The theory of planned behavior. O. rganizational Behavior and Human Decision Processes 50(2), 179–211. doi:10.1016/0749-5978(91)90020-T.

Antai, D. E., & Antai, J. B. (2008). Attitudes of women toward intimate partner violence: a study of rural women in Nigeria. Rural Remote Health 8(3), 996. https://www.ncbi.nlm.nih.gov/pubmed/18842071

Babalola, S., Kusemiju, B., Calhoun, L., Corroon, M., & Ajao, B. (2015). Factors associated with contraceptive ideation among urban men in Nigeri. Ian.t J Gynaecol Obstet, 130 Suppl 3, E42–46. 10.1016/j.ijgo.2015.05.006

Banerjee, A., La Ferrara, E., and Orozco-Olvera, V.H.,. (2019). The Entertaining Way to Behavioral Change: Fighting HIV with MTV. In W. Bank. (Ed.), Policy Research Working P.aper Washington, DC.

Bergmann, M. (2011). Ipfweight: Stata module to create adjustment weights for surveys. Retrieved November 18, 2020 from https://econpapers.repec.org/software/bocbocode/s457353.h.tm

Bott, S., Morrison, A., & World Bank. (2005). Preventing and responding to gender-based violence in middle and low-income countries a global review and analysis. World Bank,. http://econbeta.worldbank.org/external/default/main?pagePK=64165259&piPK=64165421&menuPK=64166093&theSitePK=469372&entityID=000012009_20050601161101

Callaway, B. a. S. A., Pedro H.C., (2021). Difference-in-Differences with multiple time periods. Journal of Econometrics, 225, 200-230.

Chamberlain, G. (1980). Analysis of Covariance with Qualitative Data. In N. B. o. E. Research (Ed.). Cambridge, MA.

Ellsberg, M., Jansen, H. A., Heise, L., Watts, C. H., Garcia-Moreno, C., Health, W. H. O. M.-c. S. o. W. s., & Domestic Violence against Women Study, T. (2008). Intimate partner violence and women’s physical and mental health in the WHO multi-country study on women’s health and domestic violence: an observational study. Lancet, 371(9619), 1165–1172. 10.1016/S0140-6736(08)60522-X

Fleming K. K. L. (2016). “She keeps his secrets”: A gendered analysis of the impact of shame on the non-disclosure of sexual violence in one low-income South African community. African Safety Promotion: A Journal of Injury and Violence Prevention, 11(2), 107–124.

Fontes, L. A., & Plummer, C. (2010). Cultural issues in disclosures of child sexual abuse. J Child Sex Abus, 19(5), 491–518. 10.1080/10538712.2010.512520

Gage, A. J., & Thomas, N. J. (2017). Women’s Work, Gender Roles, and Intimate Partner Violence in Nigeria. Arch Sex Behav, 46(7), 1923–1938. 10.1007/s10508-017-1023-4

Gilbert, L. K., Annor, F. B., & Kress, H. (2022). Associations Between Endorsement of Inequitable Gender Norms and Intimate Partner Violence and Sexual Risk Behaviors Among Youth in Nigeria: Violence Against Children Survey, 2014. J Interpers Violence, 37(11-12), NP8507–NP8533. 10.1177/0886260520978196

Ikuteyijo, O. O., Akinyemi, A. I., & Merten, S. (2022). Exposure to job-related violence among young female sex workers in urban slums of Southwest Nigeria. BMC Public Health, 22(1), 1021. 10.1186/s12889-022-13440-1

Ikuteyijo, O. O., Akinyemi, A.I., Merten, S., Fetters, M.D.,. (2022). Removing barriers to utilisation of support services for abused female adolescents in Nigeria slums. European Journal of Public Health, 32. 10.1093/eurpub/ckac131.455

Jina, R., & Thomas, L. S. (2013). Health consequences of sexual violence against women. Best Pract Res Clin Obstet Gynaecol, 27(1), 15–26. 10.1016/j.bpobgyn.2012.08.012

Joffe, M. M., Ten Have, T. R., Feldman, H. I., & Kimmel, S. E. (2004). Model selection, confounder control, and marginal structural models: review and new applications. The American Statistician, 58(4), 272–279.

Kolenikov, S. (2014). Calibrating survey data using iterative proportional fitting (raking). Stata Journal, 14, 22–59.

Kolenikov, S. (2019). Updatesd to the ipfraking ecosystem. Stata Journal, 19, 143–184.

Lunceford, J. K., & Davidian, M. (2004). Stratification and weighting via the propensity score in estimation of causal treatment effects: a comparative study. Statistics in medicine, 23(19), 2937–2960.

Meekers, D., Pallin, S. C., & Hutchinson, P. (2013). Prevalence and correlates of physical, psychological, and sexual intimate partner violence in Bolivia. Glob Public Health, 8(5), 588–606. 10.1080/17441692.2013.776093

National Population Comission (NPC) Nigeria, U. N., and the U.S. Centers for Disease Control and Prevention., (2016). Violence Against Children in Nigeria: Findings from a National Survey, 2014.

National Population Commission (NPC) [Nigeria], & ICF. (2019). Nigeria Demographic and Health Survey 2018. N. a. ICF.

Nguyen, K. H., Kress, H., Atuchukwu, V., Onotu, D., Swaminathan, M., Ogbanufe, O., Msungama, W., & Sumner, S. A. (2021). Disclosure of Sexual Violence Among Girls and Young Women Aged 13 to 24 Years: Results From the Violence Against Children Surveys in Nigeria and Malawi. J Interpers Violence, 36(3-4), NP2188–2204NP. 10.1177/0886260518757225

Pulerwitz, J., & Barker, G. (2007). Measuring attitudes toward gender norms among young men in Brazil: Development and psychometric evaluation of the GEM scale. Men and Masculinities, 10(3), 322–338. 10.1177/1097184X06298778

Sant’Anna, P. H. C., Zhao, Jun,. (2020). Doubly robust difference-in-differences estimators. Journal of Econometrics, 219, 101–122.

Sardinha, L., Maheu-Giroux, M., Stockl, H., Meyer, S. R., & Garcia-Moreno, C. (2022). Global, regional, and national prevalence estimates of physical or sexual, or both, intimate partner violence against women in 2018. Lancet, 399(10327), 803–813. 10.1016/S0140-6736(21)02664-7

Singhal, A., & Rogers E. M.,. (1999). Entertainment-education: A communication strategy for social change. Routledge.

Singhal, A., Cody, M. J., Rogers, E. M., & Sabido, M. (Eds.). (2004). Entertainment-education and social change. Routledge.

Singhal, A., & Rogers, E. M. (2001). The Entertainment-Education Strategy in Campaigns. In R. E. Rice & C. Atkins (Eds.), Public Communication Campaigns (3rd edition ed.). Sage Publications.

Sumner S. A., M., A.A., Saul J., Motsa-Nzuza N., Kwesigabo G., Buluma R., Kilbane T. (2015). Prevalence of sexual violence against children and use of social services-seven countries, 2007-2013. Morbidity and Mortality Weekly Report, 64(21), 565–569.

Ullman, S. E., Starzynski, L. L., Long, S. M., Mason, G. E., & Long, L. M. (2008). Exploring the relationships of women’s sexual assault disclosure, social reactions, and problem drinking. J Interpers Violence, 23(9), 1235–1257. 10.1177/0886260508314298

Umana, J. E., Fawole, O. I., & Adeoye, I. A. (2014). Prevalence and correlates of intimate partner violence towards female students of the University of Ibadan, Nigeria. BMC Womens Health, 14, 131. 10.1186/1472-6874-14-131

Usdin, S., Singhal, A., Shongwe, T., Goldstein, S., & Shabalala, A.,. (2004). No short cuts in entertainment-education: Designing Soul City step-by-step. In M. J. C. A. Singhal, E. M. Rogers & M. Sabido, (Ed.), Entertainment-education and social change: History, research, and practice (pp. 153–175). Lawrence Erlbaum Associates.

Wooldridge, J. M. (2002). Econometric analysis of cross section and panel data. MIT Press.

World Health Organization. (2021). Violence against women prevalence estimates, 2018.

World Health Organization, & Pan American Health Organization. (2012). Understanding and addressing violence against women: Intimate partner violence. https://apps.who.int/iris/bitstream/handle/10665/77432/WHO_RHR_12.36_eng.pdf

Yue Z, W. H., Singhal A.,. (2019). Using television drama as entertainment-education to tackle domestic violence in China. Journal of Development Communication, 30, 30–44.

